# Reach and public health implications of proposed new food marketing regulation in Germany: an updated analysis

**DOI:** 10.1101/2023.11.08.23298259

**Authors:** Anna Leibinger, Nicole Holliday, Oliver Huizinga, Peter von Philipsborn

## Abstract

**Introduction:** Advertising of unhealthy foods to children is a key public health concern as exposure to such advertising has been shown to adversely affect children’s food preferences, choices, purchases and intake. The proposed Children’s Food Advertising Act (Kinder-Lebensmittel-Werbegesetz or KLWG) in Germany aims to regulate such marketing by using an adapted version of the World Health Organization Regional Office for Europe Nutrient Profile Model (WHO NPM). Since the first announcement of the proposed new legislation in February 2023, several revisions to the proposed law have been made. In the present study, we evaluate the reach and public health implications of the planned law’s latest draft proposed in June 2023, updating a previously published analysis of the initial proposal.

**Methods:** Our analysis is based on a dataset from the open-source online database Open Food Facts, comprising 660 food products randomly selected from the German market. We assigned these products to the 22 food and beverage categories covered the WHO NPM. We then applied the nutrient and ingredient thresholds of the the June 2023 version of the KLWG to this sample to assess the share of products permitted for marketing to children under the proposed legislation.

**Results:** Applying the adjustments from the June 2023 KLWG version increased the proportion of products allowed for marketing to children in three product categories, namely plant-based milks (from 70% to 73%), yogurt and cream (from 13% to 73%) and fresh and frozen meat, fish and eggs (from 93% to 100%). Overall, this raised the median share of products across all 22 product categories permitted for marketing to children from 38% to 55%, with an interquartile range of 11%-73%.

**Conclusion:** Our previous study found the use of the WHO NPM, as part of the KLWG, to be feasible in the German context and suggested additional threshold adjustments. The latest draft of the KLWG from June 2023 incorporated several of these, substantially increasing the proportion of products allowed for marketing to children. This analysis provides an updated perspective on the evolving debate and its implications for public health.

## Introduction

Childhood obesity has become a pressing public health concern, in Germany and worldwide. Approximately 15% of children and adolescents aged three to seventeen in Germany, equating to nearly two million children in total, have overweight, and nearly 6% have obesity [1]. As children are exposed to marketing of unhealthy food products through various media, concerns about their physical and psychological well-being have been raised. On average, children using media in Germany are exposed to 15 food advertisements and promotions for high-sugar, high-fat, or high-salt foods daily [2]. 92% of food advertisements encountered by children in digital and television platforms in Germany are for less healthy products such as fast food, snacks, or sweets [2]. More recently, social media and influencer-based marketing campaigns have emerged as influential channels [3]. There is strong direct and indirect evidence that exposure to marketing of unhealthy foods is associated with an increased risk for childhood overweight and obesity [4-8].

Recognizing this issue, the German Federal Ministry of Food and Agriculture (BMEL), in February 2023, proposed legislation, the Children’s Food Advertising Act (Kinder-Lebensmittel-Werbegesetz or KLWG), aimed at regulating the marketing of unhealthy food products to children [9, 10]. In this draft legislation, the BMEL proposes to use an adapted version of the World Health Organization Regional Office for Europe Nutrient Profile Model (WHO NPM) in order to determine which food products should be restricted from being marketed to children [11]. The WHO NPM sets different thresholds for total fat, saturated fat, total sugars, sodium, and energy (kcal) for 22 food and beverage categories. Additionally, added sugars or artificial sweeteners rule out the marketing to children in some categories [11]. In the initial proposal for the KLWG published in February 2023, two adaptations to the original WHO NPM were included: The total sugar threshold for 100% fruit juice includeded in the original WHO NPM was removed, as was the total fat threshold for dairy milk drinks [12, 9].

Further adaptations to the WHO NPM were introduced in a later draft of the proposed legislation dating from June 2023 [13]. In particular, the total fat threshold for plant-based milks, and the total fat and saturated fat thresholds for yogurt, sour milk, cream and similar foods were removed, as was the total fat threshold for fresh and frozen meat, poultry, fish and similar products. Restrictions regarding advertising time and location have also been loosened in this latest draft [13]. In the present paper, we analyze how these adaptations affect the share of products permitted for marketing to children, updating an earlier analysis based on the initial draft for the KLWG [14].

## Methods

We used the same dataset that we used in our previous study [14]. For this, we randomly sampled food products on the German market from the open-source online database Open Food Facts. Subsequently, we followed the guidelines outlined in the WHO NPM manual to systematically assign these products to the 22 product categories specified by the WHO NPM [11]. We randomly selected 30 products for each category, totaling 660 products in total. Using the resulting data set we applied the WHO NPM thresholds with the adjustments proposed in the June 2023 version of the KLWG to determine which products would be below the thresholds and would therefore be allowed for marketing to children. We did this for all 22 food categories to derive the share of products that would be allowed for marketing to children for each category. We then calculated the median share and its interquartile range of products that would be permitted for marketing to children across all 22 categories. A detailed description of our methodology is included in the publication presenting the results of our analysis based on the February 2023 draft [14].

All analyses were carried out using R. To visually represent our findings, we utilized Microsoft Excel’s conditional formatting feature to generate tables with color-coded elements. Within this color scheme, various shades of green signify favorable outcomes from a public health standpoint, indicating a high proportion of products that do not exceed nutritient or ingredient thresholds. Conversely, shades of red represent less favorable results, while shades of yellow denote values that fall in between.

## Results

Applying the adaptions from the June 2023 version of the KLWG to our dataset raised the share of products that would be permitted for marketing to children from 70% to 73% in the category of plant-based milks, from 13% to 73% in the category of yogurt, sour milk, cream and similar foods, and from 93% to 100% in the category of fresh and frozen meat, poultry, fish and similar. Overall, this raised the median share of products that would be allowed for marketing to children across the 22 product categories of the WHO NPM from 38% to 55% (IQR: 11%-73%) (see Table 1).

**Table 1:** Share of products meeting all nutrient and ingredient criteria of the WHO NPM (i.e., permitted for marketing to children)

|  | Original WHO NPM | WHO NPM with adaptations of first draft (Feb 2023)* | WHO NPM with adaptations of latest draft (June 2023)** |
| --- | --- | --- | --- |
| **Median across product categories (IQR)** | **20%**<br>**(3%-59%)** | **38%**<br>**(11%-73%)** | **55%**<br>**(11%-73%)** |
| **1 Confectionery** | 0% | 0% | 0% |
| **2 Cakes and cookies** | 0% | 0% | 0% |
| **3 Savoury snacks, nuts and seeds** | 53% | 53% | 53% |
| **4.1 Juices** | 0% | 100% | 100% |
| **4.2 Dairy milk drinks** | 20% | 80% | 80% |
| **4.3 Plant-based milks** | 70% | 70% | 73% |
| **4.4 Energy drinks** | 0% | 0% | 0% |
| **4.5 Soft drinks, bottled water and other drinks** | 23% | 23% | 23% |
| **5 Ice cream** | 0% | 0% | 0% |
| **6 Breakfast cereals** | 57% | 57% | 57% |
| **7 Yogurt and cream** | 13% | 13% | 73% |
| **8 Cheese** | 20% | 20% | 20% |
| **9 Ready-made and convenience foods** | 60% | 60% | 60% |
| **10 Butter, other fats and oils** | 73% | 73% | 73% |
| **11 Bread** | 57% | 57% | 57% |
| **12 Pasta and grains** | 93% | 93% | 93% |
| **13 Fresh and frozen meat, fish and eggs** | 93% | 93% | 100% |
| **14 Processed meat and fish** | 13% | 13% | 13% |
| **15 Fresh and frozen fruit and vegetables** | 100% | 100% | 100% |
| **16 Processed fruit and vegetables** | 20% | 20% | 20% |
| **17 Savoury plant-based foods** | 10% | 10% | 10% |
| **18 Sauces, dips and dressings** | 0% | 0% | 0% |

Abbreviations: WHO NPM: World Health Organization Regional Office for Europe Nutrient Profile Model; IQR: interquartile range, shown here as span from the 25th to the 75th percentile. \*This is the original WHO NPM with two adaptations proposed by Germany's Federal Ministry of Food and Agriculture in the original draft (February 2023) of the proposed law, namely a removal of the total sugar threshold for 100% fruit juice, and of the total fat threshold for milk. \*\*This is the original WHO NPM with adaptations proposed by Germany's Federal Ministry of Food and Agriculture in the latest draft (June 2023) of the proposed law, namely a removal of the total sugar threshold for 100% fruit juice, of the total fat threshold for milk, of the total fat threshold for plant-based milks, of the total fat and saturated fat threshold for yogurt and cream, and of the total fat threshold for fresh and frozen meat, fish and eggs.

## Discussion

The proposed new food marketing legislation in Germany has sparked a lively public debate. In our initial analysis, we found the use of the WHO NPM as part of the KLWG to be feasible and appropriate in light of the public health objective of limiting children’s exposure to marketing for products high in salt, sugar and fat [14]. We also found that a substantial share of products on the German market met the nutrient and ingredient thresholds defined by the WHO NPM, and would still be permitted for marketing to children under the proposed legislation [14]. The adaptations to the WHO NPM introduced in the proposed law’s latest draft further increase the share of products permitted for marketing to children to a median of 55% across the 22 product categories of the WHO NPM. This suggests that the proposed law meets its objective of limiting marketing specifically for less healthy products, as defined by the evidence-informed nutrient and ingredient thresholds of the adapted WHO NPM.

## Data Availability

All data produced in the present study are available upon reasonable request to the authors

https://doi.org/10.17605/OSF.IO/BJEVC

## Statements

### Statement of Ethics

No human subjects were involved in this research and no ethical clearance was sought in line with the regulations of the Ethics Committee of Ludwig-Maximilians-Universität München (LMU Munich).

### Conflict of Interest Statement

PvP reports receiving research funding from Germany’s Federal Ministries of Food and Agriculture (BMEL), Education and Research (BMBF) and the Environment and Consumer Protection (BMUV), as well as travel costs and speaker and manuscript fees from the German and Austrian Nutrition Societies (DGE and ÖGE), the German Diabetes Society (DDG) and the German Obesity Society (DAG). OH is an employee of the German Diabetes Society (DDG) and the German Obesity Society (DAG), and has previously been an employee of foodwatch. The staff positions of AL, NH and PvP are funded through research grants from Germany’s Federal Ministries of Food and Agriculture (BMEL) and Education and Research (BMBF).

### Funding Sources

This work was conducted without external funding through staff positions at Ludwig-Maximilians-Universität München (LMU Munich), Germany, and the German Obesity Society (DAG), Berlin, Germany.

### Author Contributions

Conceptualization: PvP and OH; Methodology: PvP, OH, NH, AL; Investigation: AL, NH, PvP. Data Curation: NH and AL; Formal Analysis: AL and NH; Writing – Original Draft: AL; Writing – Review and Editing: PvP, NH, AL and OH; Supervision: PvP.

### Data Availability Statement

The data that support the findings of this study are openly available on the Open Science Framework at https://doi.org/10.17605/OSF.IO/BJEVC [15]

## Notes

### Clinical Protocols

https://doi.org/10.17605/OSF.IO/BJEVC

### Funding Statement

This work was conducted without external funding through staff positions at Ludwig-Maximilians-Universitaet Muenchen (LMU Munich), Germany, and the German Obesity Society (DAG), Berlin, Germany.

